# Excess deaths reveal the true spatial, temporal, and demographic impact of COVID-19 on mortality in Ecuador

**DOI:** 10.1101/2021.02.25.21252481

**Authors:** Leticia Cuéllar, Irene Torres, Ethan Romero-Severson, Riya Mahesh, Nathaniel Ortega, Sarah Pungitore, Nicolas Hengartner, Ruian Ke

**Author notes:** Correspondence should be addressed to: Leticia Cuéllar, Irene Torres, Nicolas Hengartner, Ruian Ke.

## Abstract

**Background:** In early 2020, Ecuador reported one of the highest surges of per capita deaths across the globe.

**Methods:** We collected a comprehensive dataset containing individual death records between 2015 and 2020 from the Ecuadorian National Institute of Statistics and Census and the Ecuadorian Ministry of Government. We computed the number of excess deaths across time, geographical locations and demographic groups using Poisson regression methods.

**Results:** Between January 1^st^ and September 23^rd^, 2020, the number of excess deaths in Ecuador is 36,402 (95% CI: 35,762-36,827) or 208 per 10^5^ population, which is 171% of the expected deaths in that period in a typical year. Only 20% of the excess deaths are attributable to confirmed COVID-19 deaths. Strikingly, in provinces that were most affected by COVID-19, such as Guayas and Santa Elena, the all-cause deaths are more than double the expected number of deaths that would have occurred in a normal year. The extent of excess deaths in men is higher than in women, and the number of excess deaths increases with age. Indigenous populations had the highest level of excess deaths among all ethnic groups.

**Conclusions:** Overall, the exceptionally high level of excess deaths in Ecuador highlights the enormous burden and heterogeneous impact of COVID-19 on mortality especially in older age groups and indigenous populations in Ecuador that was not fully revealed by COVID-19 death counts. Together with the limited testing in Ecuador, our results suggest that the majority of the excess deaths were likely to be undocumented COVID-19 deaths.

## Introduction

The coronavirus disease 2019 (COVID-19) pandemic has caused high morbidity and mortality across the globe. The total number of confirmed COVID-19 deaths is approximately 1.8 million by the end of 2020^1^. However, this number only partially reflects the total burden of COVID-19 on mortality, because it does not include undocumented COVID-19 deaths or other causes of deaths due to societal disruptions. A more accurate measure of the burden of COVID-19 on mortality is therefore the excess deaths^2, 3^, i.e. the number of all-cause deaths exceeding the expected number of deaths in a typical year without COVID-19. This measure is especially relevant for countries where testing capacity is limited, such as Ecuador^4^, which may lead to substantial underreporting of COVID-19 cases and deaths.

Excess deaths in 2020 have been formally estimated previously for many high-income countries with high COVID-19 cases, including the United States (US)^5-8^, England and Wales^9^, Italy^10, 11^, and middle income countries, such as Brazil^12^. The numbers of all-cause deaths range between 120% and 131% of expected deaths in these countries, i.e. between 20% and 31% of excess deaths, highlighting the heavy burden of COVID-19 spread directly on mortality, despite differences in demographics, social mixing patterns and health care systems. In these countries, a major fraction of excess deaths (between 67% to 80%) are attributable to COVID-19 deaths^5, 8, 9,11^, suggesting that the majority of excess deaths are caused directly by COVID-19 infections. In contrast, Ecuador reported a relatively small number of COVID-19 deaths^13^; however, a surprisingly large number of excess deaths was reported in the Our World in Data online database^13^, in a newspaper during early COVID-19 outbreak in April 2020^14^ and more recently in a study^15^. This raises the questions about how the excess deaths vary temporally, spatially and demographically and how they are related to COVID-19 spread in Ecuador.

Here, by analysing a rich dataset containing death records from Ecuador during 2015-2020, we provide a comprehensive analysis of the spatial, temporal and demographic patterns of excess all-cause deaths in Ecuador between January 1 and September 23 of 2020. We show that the extent of excess deaths in Ecuador is several folds higher than many countries severely affected by COVID-19. In addition, we found the spatial, temporal and demographic pattern of excess deaths reveal a much more severe early burden of COVID-19 in Ecuador and a more complex spaciotemporal distribution of COVID-19 deaths than is apparent in the official death reports.

## Methods

### Data

Death records from 2015-2019 for all-cause mortality were obtained from the Ecuadorian National Institute of Statistics and Census. The records include age, sex, and ethnicity of the diseased, place of death registration, residence, and the International Classification of Disease (ICD) code for the cause of death. The Ecuadorian Ministry of Government provided death records from January 1st, 2020 to September 26th, 2020 containing sex, age, and registration and residence location by parish, canton, and province, but without the cause of death. There are 24 provinces in Ecuador. We ignore the three smaller “Not Delimited Areas” located along various provinces’ borders. We use the 2020 population estimates from the INEC (the Ecuadorian National Institute of Statistics and Census).

Individual records of COVID-19 incidence and testing for Ecuador were obtained from the Ecuadorian Ministry of Public Health. All records were aggregated at the weekly level and binned by sex, age group, and province. Ethics committee approval was obtained from the Ecuadorian Ministry of Public Health. The analysis by Los Alamos National Laboratory team was approved by that institution’s IRB.

### Statistical Methodology

To estimate the expected number of deaths in the absence of COVID-19, we fit a Poisson regression to the binned weekly death counts from 2015 to 2019. The regression predicts the number of weekly deaths as a function of the week of the year to account for annual variations, the province, and deceased demographics (sex and age group). Since the 2020 death records have ethnicity up to July 30, we fitted a second model adding ethnicity as a covariate.

We calculate the number of weekly excess deaths in 2020, by subtracting from the 2020 deaths the number of expected or baseline deaths calculated with the Poisson regression model. We define the excess death factor (EDF) as the ratio of 2020 observed deaths over the expected deaths. See Supplementary Materials for details of the statistical model and the calculation of 95% confidence intervals for the number of deaths.

## Results

### Excess deaths at the country level

We first predicted the expected number of all-cause deaths between January 1 and September 23, 2020 using a Poisson regression model fitted to deaths data collected between 2015 and 2019 (Fig. S1 and see Methods and Supplementary Materials).We estimated that the expected number of all-cause deaths up to September 23, 2020 is 51,360 (CI: 50,935-52,000). The number of reported total all-cause deaths during this period is 87,762, leading to the number of excess deaths of 36,402, i.e. 71% higher than expected (Table 1). This is in stark contrast to the percentages estimated for the US (20% of excess deaths)^5, 8^, England and Wales (31%)^9^, and Brazil (22%)^12^. Normalized by the population size of Ecuador in 2020 (i.e. 17,468,736 according to the INEC (the Ecuadorian National Institute of Statistics and Census), the number of total all-cause deaths is 502 per 10^5^ population. The number of excess deaths is 208 per 10^5^ population, i.e. almost 3 fold higher than the estimate for the US (72 per 10^5^ population)^5^.

**Table 1.**
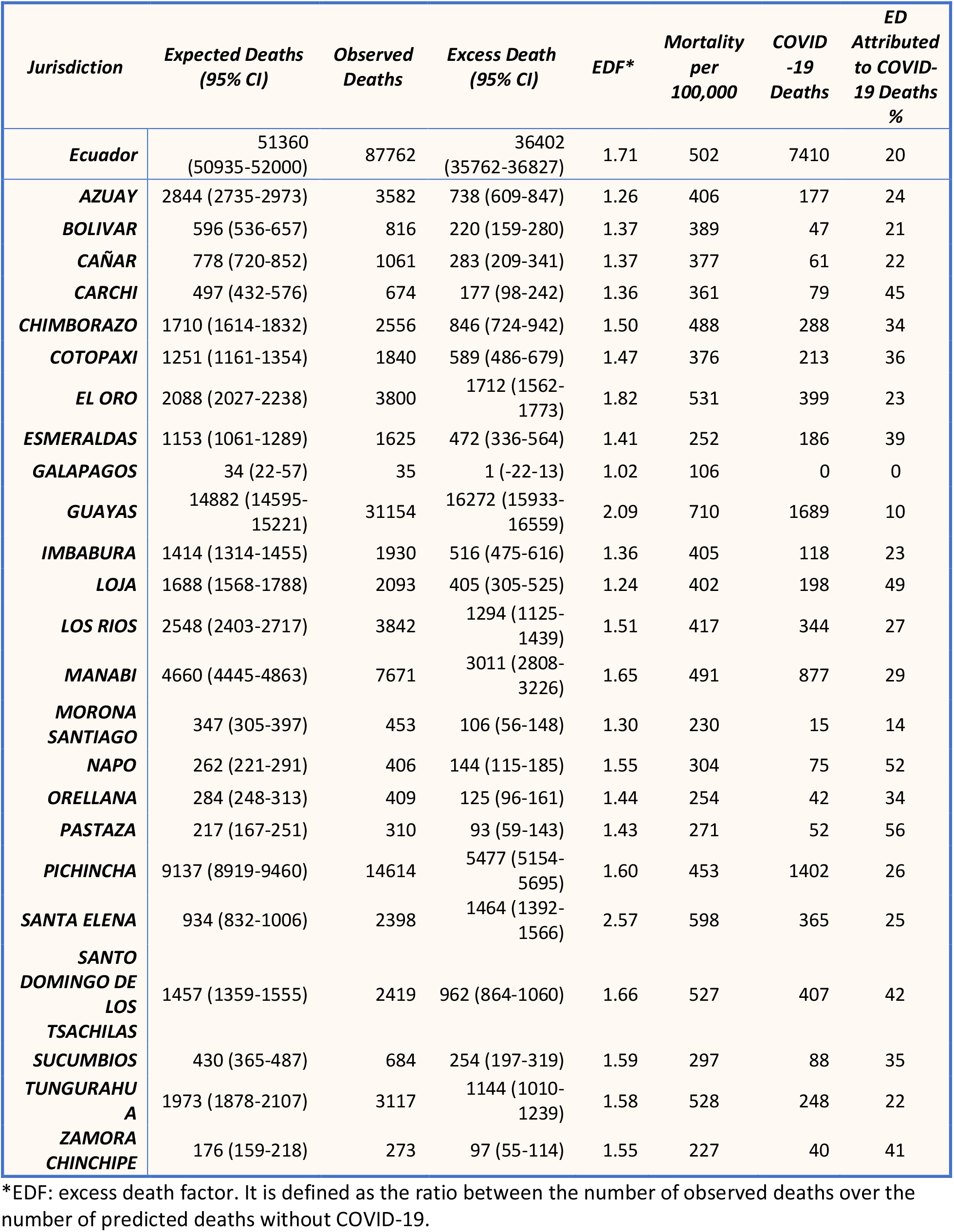
Mortality and estimated excess deaths between January 1 and September 23, 2020.

Up to September 23, 2020, there were 7,410 officially reported COVID-19 deaths in Ecuador, which only account for 20% of the total all-cause excess deaths (36,402). This is a much lower percentage than those reported in countries, such as the US (67%)^5,8^, and England and Wales (87%)^9^.

Time series of the total all-cause deaths suggest that the number of reported deaths started to become higher than expected in mid-March 2020, shortly after the first COVID-19 case was confirmed in Ecuador on February 29 (Fig. 1). There exist two waves of excess deaths, i.e. a first major wave between late March and April, and a second minor wave between July and early August (Fig. 1). The first wave started in the week of March 11-17. The number of excess deaths increased extremely rapidly and reached the highest level, i.e. 33 per 10^5^ population per week (or a total of 7,133 deaths per week; see Fig. S2) in the first week of April. Remarkably, this is over 5 times the number of expected deaths per week (Fig. S2). The number of weekly excess deaths declined afterwards to approximately 5 per 10^5^ population per week (Fig. S2). This rapid decline coincides with the period of strict lockdown implemented by the Ecuador government^16^.

**Figure 1.**
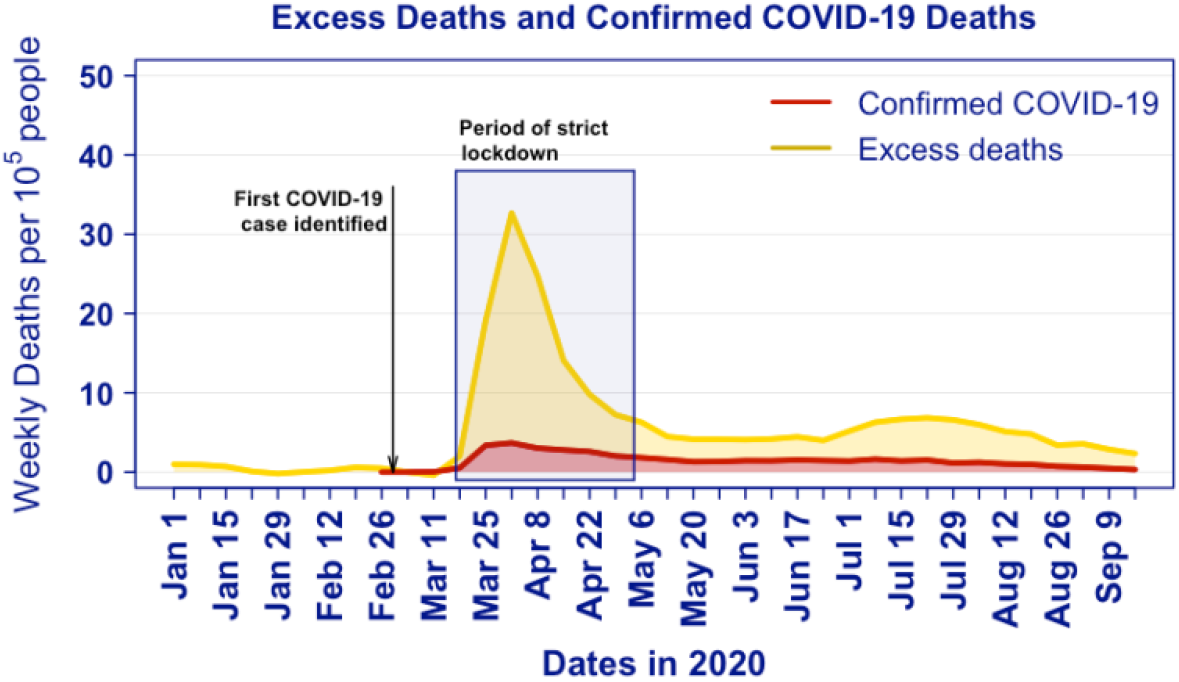
Time series for excess deaths (yellow) and documented COVID-19 deaths (red) per 100,000 people in Ecuador. The period of strict national lockdown by the Ecuador government (shaded area) is taken from the Oxford COVID-19 Government Response Tracker^16^.

The second wave occurred in July and early August, a period after strict lockdowns were gradually phased out. The number of weekly per capita deaths ranges between 5 and 8 per 10^5^ population or total weekly deaths between 2,200 and 2,500 (Fig. 1 and S2). These numbers are between 170% and 190% of the expected number of deaths.

### Excess deaths by sex and age and ethnicity

We estimate that over the entire observational period, excess deaths were higher in men than women, i.e. 271 and 147 excess deaths per 10^5^ people for men and women, respectively (Fig. 2A). The observed death in men was 183% of expected levels, compared to 156% for women (Fig. S3A).Deaths was significantly elevated for all age groups greater than 40 years, with the number exponentially increasing with age group (Fig. 2B). For example, the number of excess deaths is 93 per 10^5^ people for the age group between 40 and 49, and increases to 4,428 per 10^5^ people for the age group older than 80. Across all age groups greater than 40 years old, more men died than women (Fig. 2C).

**Figure 2.**
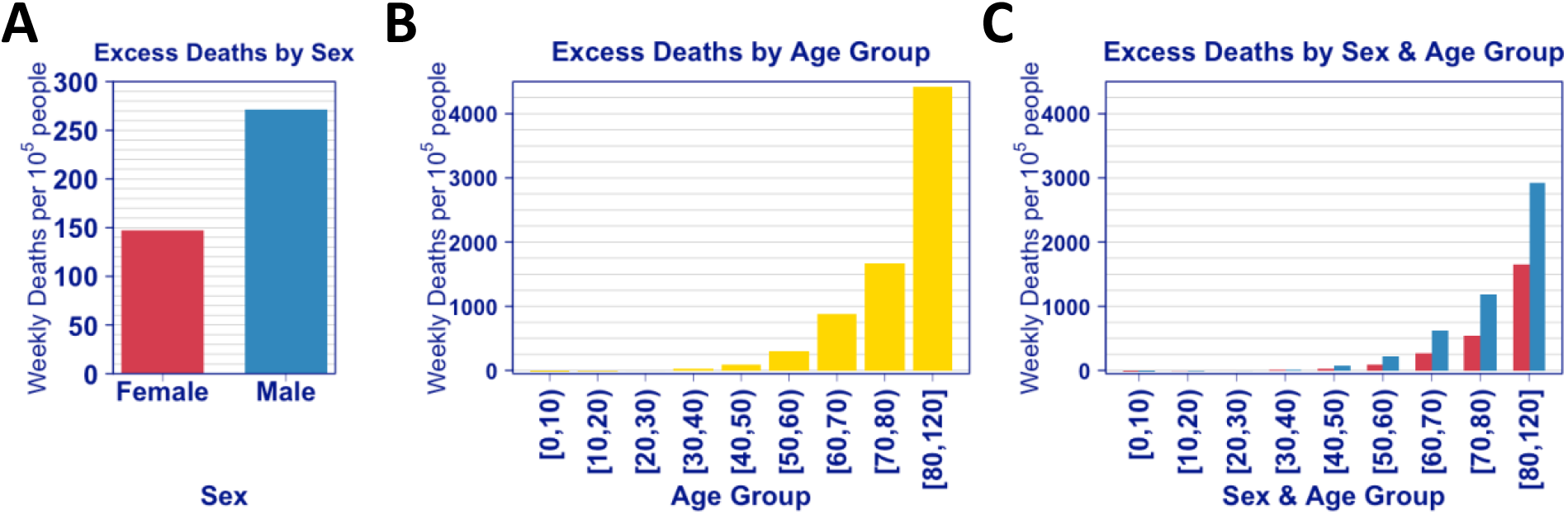
Excess deaths per 100,000 people by sex (A), by age group (B) and by sex and age group (C) in Ecuador. The numbers of excess deaths are normalized by the population size in each group.

Interestingly, we found that the numbers of deaths were 27% and 22% lower than expected for age groups between 0 and 9 years and between 10 and 19 years (Fig. S3B). A slight decrease of mortality in children has been observed in England^17^. One possible explanation is that the social distancing and restrictions during the pandemic may be protective for children from accidental deaths.

The sex and age characteristics in excess deaths are in line with the demographic characteristics of the risk of COVID-19 mortality^18^. The fact that the number of reported COVID-19 deaths only account for 20% of the total all-cause deaths suggests that a large fraction of COVID-19 deaths are undocumented, as a result of lack of COVID-19 testing^4^. To corroborate this hypothesis, we obtained COVID-19 testing and diagnoses data from the Ministry of Public Health (Methods), and calculated the weekly testing positivity rate, i.e. the number of COVID-19 cases over the total number of tests administered. The overall testing positivity rates in Ecuador using data up to September 23 is exceptionally high, i.e. 35%. In particular, during the period between March 18^th^ and April 8^th^ when highest excess deaths and COVID-19 deaths are observed, the test-positivity rates are as high as 37-47% (Fig. S4). These rates are in stark contrast to the test positivity rate reported in other countries^13^. For example, in the US and the United Kingdom, where testing has been limited during the early period of COVID-19 outbreak, the test-positivity rates were between 20-30% in April, 2020 and dropped rapidly to below 10% since early May^13^. Therefore, the exceptionally high test-positivity rates in Ecuador are consistent with the hypothesis that the majority of excess deaths in Ecuador were caused directly by COVID-19 infection, but were not documented as COVID-19 deaths as a result of limited testing capacity.

We next analysed the excess deaths by ethnic groups in Ecuador, and found that the excess death factor for the indigenous group is 2.2, i.e. excess deaths are 220% of the expected deaths compared to just 36% for the predominant ethnic group in Ecuador, i.e. mestizo group (Fig. 3A). The distribution of excess deaths by sex and age-group for the indigenous population is similar to the ones from the general population, except that excess death factors for females in the age-groups [20-30), [30-40), and [40-50), are larger than those from the corresponding male populations by 75%, 32% and 17% respectively (Fig. 3B). Ethnicity for death registrations is self-reported with typically 5% not being reported, but 25% not reported in 2020. Even if all unreported ethnic data was not indigenous, our results would show that the indigenous group is much more disproportionally affected by COVID-19 than the confirmed death counts suggest.

**Figure 3.**
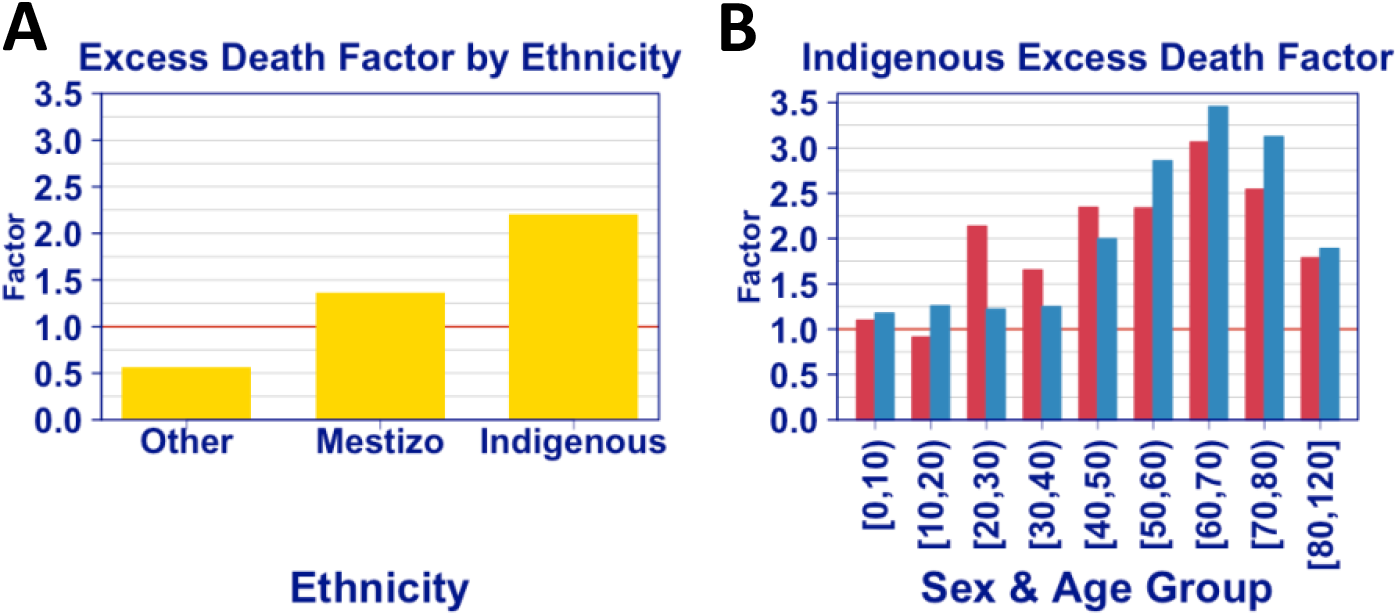
The excess death factor, i.e. the number of all-cause deaths over the number of expected deaths, by ethnicity (A) and by sex and age for the indigenous group (B).

### Spatial and temporal pattern of excess deaths at the provincial level

Next, we examined the excess deaths at the provincial level. The highest total excess deaths occurred in Guayas and Pichincha, comprising of 29% and 18% of the excess deaths in Ecuador, respectively (Table 1). Note that the two provinces also reported the highest numbers of COVID-19 deaths (Table 1). When the number of deaths is normalized by the population size in each province, Guayas and Santa Elena had the highest per capita deaths, at 710 and 598 per 10^5^ people, respectively (Table 1). Remarkably, the estimated excess death factors, i.e. the number of observed deaths over the number of expected deaths for Guayas, Pichincha and Santa Elena are 2.09, 1.60 and 2.57, respectively (Table 1). These exceptionally high levels of excess deaths rate emphasize the enormous burden of COVID-19 spread on mortality.

There is a strong spatial and temporal pattern in the magnitude and timing of excess deaths that are mostly consistent with the COVID-19 spread (as measured by COVID-19 deaths). In Guayas and Santa Elena, most excess death occurred during late-March and mid-May, 2020, with the highest numbers of weekly all-cause deaths and documented COVID-19 deaths occurred during the two weeks between April 1^st^ and 14^th^ (Fig. 4), leading to the first wave of all-cause deaths in Ecuador as seen in Fig. 1. During this period, the test-positivity rates in these provinces are extremely high (between 40-80%; see Fig. S4). Again, this highlights the burden of COVID-19 spread on the limited testing and health care capacity in Ecuador, and the notion that it is likely that a large fraction of COVID-19 deaths was not diagnosed or documented. The high burden of mortality is also apparent in provinces neighbouring Guayas and Santa Elena, such as Los Rios, Manabí, Bolivar, Chimborazo, Cañar and El Oro (Fig. 4D, S5 and S6).

**Figure 4.**
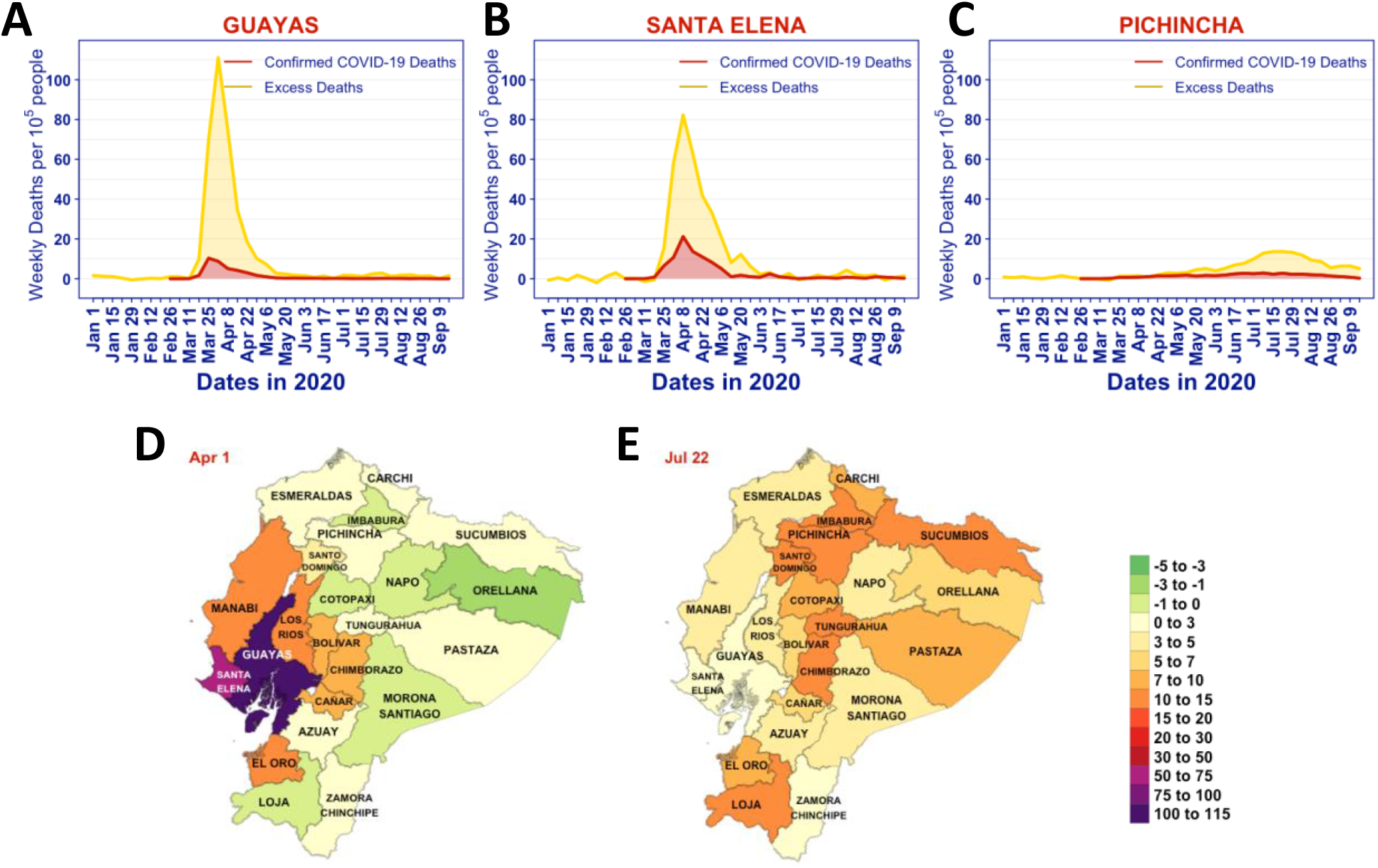
The spatial and temporal patterns of excess deaths in 2020 in Ecuador. **(A-C)** Time series for excess deaths (yellow) and documented COVID-19 deaths (red) per 100,000 people in Guayas, Santa Elena and Pichincha.**(D and E)**Provincial maps of Ecuador showing the number of excess deaths per 10^5^ population (colour) during the weeks of Apr 1-7 (panel D), and July 22-28, 2020 (panel E).

In July and August, Pichincha, where the capital city, Quito, is located, became the COVID-19 epicentre during the second wave of excess deaths (Fig. 4C and 4E). Several other provinces close to Pichincha also have seen excess deaths peaked during this period. Again, the geographical and temporal trend in the excess deaths follows the spatial and temporal spread of COVID-19, as indicated by reported COVID-19 deaths (Fig. S5 and S6). For example, the provinces that had highest total deaths also had the highest number of reported COVID-19 deaths.

## Discussion

We observed a very high level of total all-cause deaths, 87,762 in total or 502 per 10^5^ people between January 1, 2020 and September 23, 2020 in Ecuador. Analysing death records during 2015 and 2020, we estimated the number of excess deaths to be 36,402 or 208 per 10^5^ population. This is 171% of the expected number of deaths in a typical year and 71% of excess deaths. Comparing these numbers to estimates for other countries that have been heavily affected by COVID-19, such as the US, England and Wales and Brazil, we found that the level of excess deaths in Ecuador is approximately 2.9 and 2.3 folds of the estimate in the US and in England and Wales, respectively^5, 9^, highlighting the enormous burden of COVID-19 on mortality in Ecuador.

The high level of excess deaths in Ecuador was reported previously in an online database^13^, newspapers^14^ and a research study^15^. These works were based on published numbers of all-cause deaths in Ecuador. Here, we provide analyses of a comprehensive set of individual death records between 2015 and 2020 with both geographical, temporal and demographic information. By fully accounting for spatial, temporal, geographical and demographic variations in the deaths in Ecuador over 6 years, our statistical model accurately estimates how the burden of excess deaths varies across time, geographical areas and demographic groups as we discuss below.

A distinguishing feature of excess deaths in Ecuador as compared to other countries, such as the US and England and Wales^5, 8, 9^ is that COVID-19 deaths (7,410 in total up to September 23, 2020) only account for a small fraction, i.e. 20%, of all-cause excess deaths. Excess deaths can be caused directly by COVID-19 infection or indirectly as a result of COVID-19 related disruptions in society^2, 3^. We do not have data on the causes of deaths in 2020, and thus are unable to assess the major cause of the large number of excess deaths not attributable to COVID-19 deaths. However, results of our analyses as listed below strongly suggest that the true number of COVID-19 deaths is underreported and a majority of excess deaths are likely to be caused directly by COVID-19, as seen in other countries^8-11,12^. First, the spatial and temporal patterns of excess deaths follow closely the spatial and temporal patterns of COVID-19 outbreaks in Ecuador as suggested by the COVID-19 deaths counts. For example, high excess deaths occurred during periods when the number of COVID-19 deaths peaked in provinces most affected by COVID-19, such as Guayas, Santa Elena and Pichincha. Second, more men died than women, and the excess deaths become apparent for age groups greater than 40 years old and the number of excess deaths increases dramatically as age increases. These demographic characteristics of mortality are consistent with the risk of COVID-19 mortality^18^ and are very similar to the demographics in excess deaths found in countries, such as England and Wales^9^ and Italy^11^, where the majority of excess deaths were attributable to COVID-19 deaths. Third, we found that the test-positivity rate in Ecuador, is extremely high over the period of COVID-19 outbreak, indicating the very low testing capacity in Ecuador^4^. Therefore, it is likely that a lot of COVID-19 infections as well as deaths are not detected or documented. Indeed, anecdote evidence that people dying of pneumonia before being able to make to hospitals was reported during the early COVID-19 outbreak in Ecuador^14^. Overall, irrespective of the cause of the excess deaths, our analysis demonstrates the enormous burden of COVID-19 on mortality in Ecuador that is not revealed by the numbers of COVID-19 cases and deaths previously.

Non-pharmaceutical interventions played important roles in suppressing SARS-CoV-2 transmission and preventing deaths^19^. We found that in two provinces that had the most per capita excess deaths, i.e. Guayas and Santa Elena, the numbers of excess deaths decreased rapidly after the highest excess deaths occurred in the week of April 1-8. This rapid decline coincides with the strict social distancing measures implemented during March 17 and May 4, 2020 by the Ecuador government. These measures include a national lockdown, boarder closure, suspension of travel and strict confinement of citizens with a curfew^16^. Therefore, it is likely that the non-pharmaceutical interventions lead to reduced COVID-19 transmission and effectively averted the extremely rapid increases in deaths, as shown for COVID-19 outbreaks in other countries^19^. Note that the total all-cause deaths in Guayas and Santa Elena are extremely high, at 710 and 598 per 10^5^ population, corresponding to 209% and 257% of the numbers of expected deaths, respectively. Given the exponential growth nature of the outbreak in the absence of intervention efforts^20-22^, we reason that COVID-19 may have caused even more mortality than the observed high level if strict social distancing measures had been delayed or not been implemented at all. Thus, our results strongly suggest the importance of social distancing efforts in preventing deaths, and the potential devastating consequence in mortality if the strict non-pharmaceutical interventions especially during early COVID-19 outbreak were not implemented.

## Data Availability

The data underlying this article cannot be shared publicly due to Ecuador government regulations that the Ecuadorian Ministry of Public Health must approve the research protocol to release reusable COVID-19 datasets. The data may be shared on reasonable request to the corresponding authors and the Ecuadorian Civil Registry.

## Ethics approval

Ethics committee approval was obtained from the Ecuadorian Ministry of Public Health. The analysis by Los Alamos National Laboratory team was approved by that institution’s IRB.

## Author contributions

Conceptualisation: LC, IT, ERS, NH and RK; data acquisition: IT; methodology and formal analysis: LC, ERS, RM, NO, SP and NH; funding acquisition: LC and RK; underlying data validation: LC, IT and NH; visualisation and writing: LC and RK.

