## Supplementary Materials for "Excess deaths reveal the true spatial, temporal, and demographic impact of COVID-19 on mortality in Ecuador"

### Affiliations:
[1] A-1 Information Systems and Modeling, Analytics, Intelligence and Technology Division, Los Alamos National Laboratory, NM87545, USA.
[2] Fundación Octaedro, Quito, Ecuador.
[3] T-6 Theoretical Biology and Biophysics, Theoretical Division, Los Alamos National Laboratory, NM87545, USA.
[4] Department of Biology, University of Texas, Austin, Texas, 78712, USA.
[5] University of California Santa Barbara, Santa Barbara, CA 93106, USA.
[6] Program in Applied Mathematics, University of Arizona, Tucson, Arizona 85721, USA.

### Supplementary Methods

#### Statistical Model

To estimate the number expected deaths in 2020 (without COVID-19), we model the number of weekly deaths as a Poisson distributed random variable with expected value $\lambda_{Bijkl}$ and assume that the logarithm of its expected value can be expressed as a linear combination of a set of explanatory variables

$$\log(\lambda_{ijkl}) = c_0 + c_{1,i}Week_i + c_{2,j}\,Province_j + c_{3,k}Sex_k + c_{4,l}Age\_Group_l + c_{5,s}\,Sex_k * Age\_Group_l$$

where $\lambda_{ijkl}$ is the expected number of deaths during week $i$, in province $j$ for the sex k and age group $i$.

We estimate the parameters of this generalized linear model using the *glm* function in R (version 3.6.1), specifying family='Poisson'. We calculate the number of expected deaths per week, province, sex, and age group and aggregate the results at the province and national levels, per week, and for the different demographic groups.

We estimate 95% confidence intervals by sampling from the distribution of the estimated coefficients, a multivariate normal distribution with mean the vector of estimated parameters and covariance matrix calculated with the R "vcov" function. We use 100 thousand samples to compute a distribution for the estimated expected number of deaths for combination of week, province, sex, and age group and aggregate the estimated deaths at the province and national levels. Finally, we calculate the 2.5 and 97.5 percentiles.

The estimated model fitted with data from 2015 to 2019, fits fairly well as demonstrated by a closer inspection of the standardized residuals: square root of the observed deaths – the square root of the expected deaths. Taking the square root of the observed and the expected deaths is known as the variance stabilizing transformation that is commonly used for Poisson regression to alleviate heteroskedasticity. All estimated parameters are significant, including the interaction term between sex and age group.

#### Supplementary References

1. Hale, Thomas, Webster S, Petherick A, Phillips T, Kira B. Oxford COVID-19 Government Response Tracker, Blavatnik School of Government. . *Data use policy: Creative Commons Attribution CC BY standard* 2020.

### Supplementary Figures

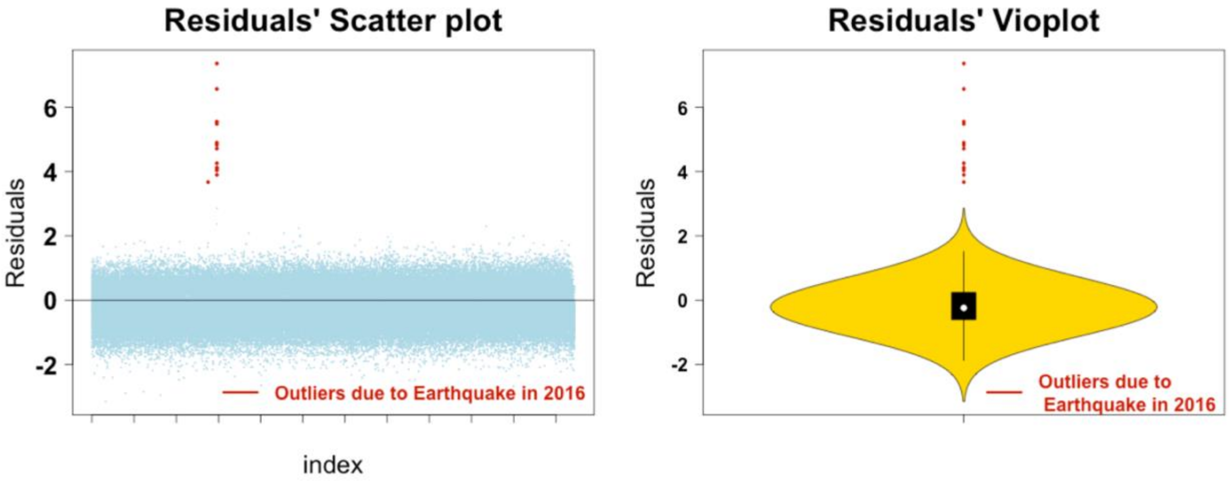


**Figure S1. Plots of standardized residuals from the Poisson regression for excess deaths.** Left plot shows scatter plot of the residuals and the right plot shows their distribution as depicted by a vioplot. Outliers (red), are cases that did not fit well the estimated model, correspond to deaths caused by the 2016 earthquake with epicentre in the province of Esmeraldas and a magnitude of 7.8 in the Mercalli intensity scale.

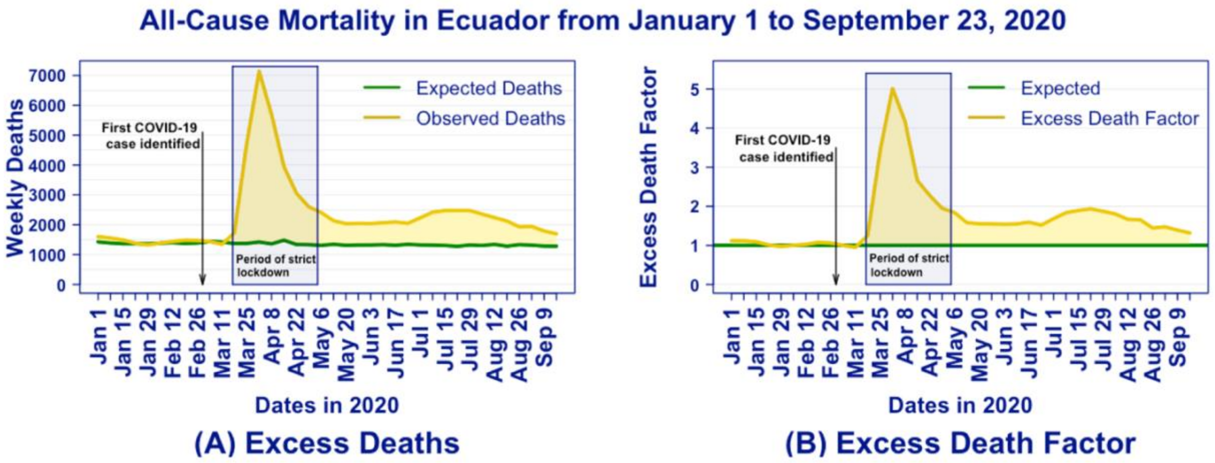


**Figure S2. Time series for (A) weekly expected deaths and observed deaths and (B) corresponding time series of excess death factor.** The period of strict national lockdown by the Ecuador government (shaded area) is taken from the Oxford COVID-19 Government Response Tracker[1].

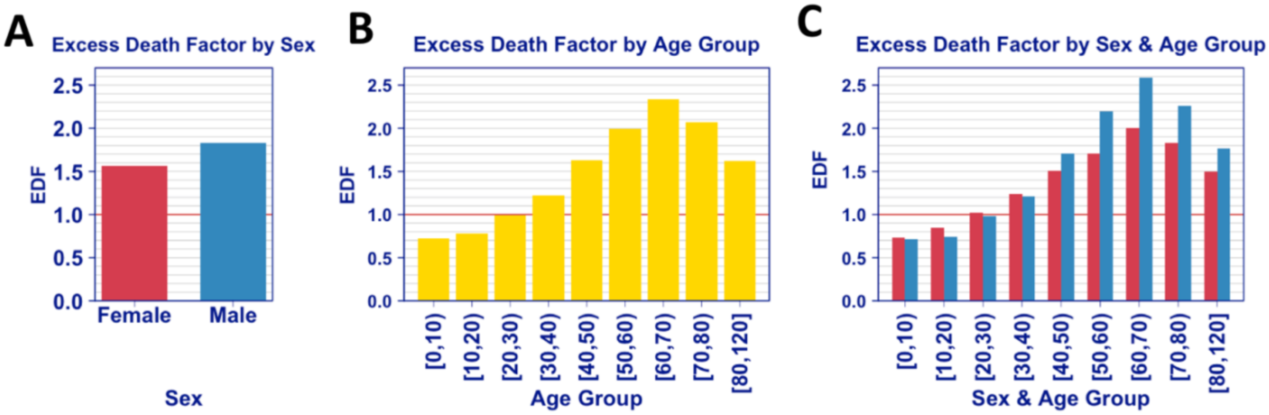


**Figure S3. The excess death factor by demographics, i.e. the number of all-cause deaths over the number of expected deaths: (A) by sex, (B) by age group, and (C) by sex and age group.** Note that the age group [60, 70) has the largest excess death factor.

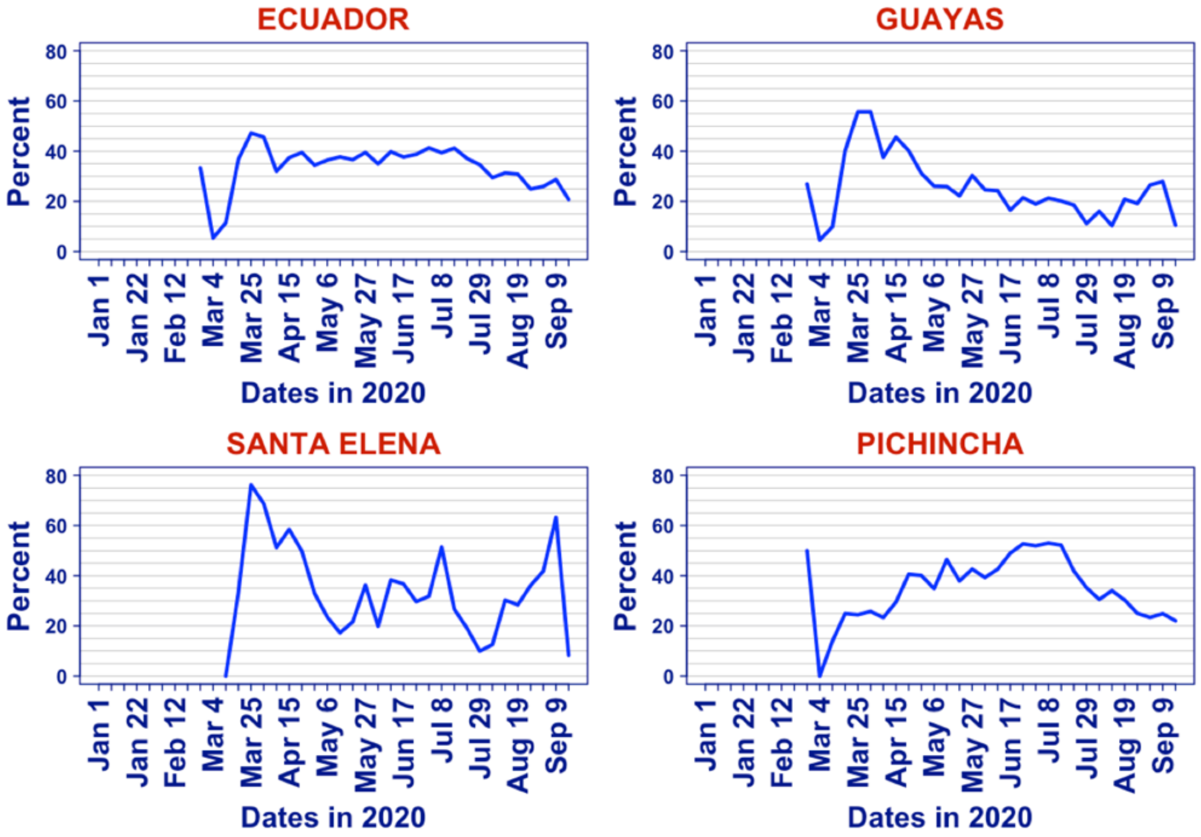


**Figure S4. Time series of COVID-19 test positivity rate, i.e. percent of positive COVID-19 tests for Ecuador (national level), and for the provinces of Guayas, Santa Elena, and Pichincha.**

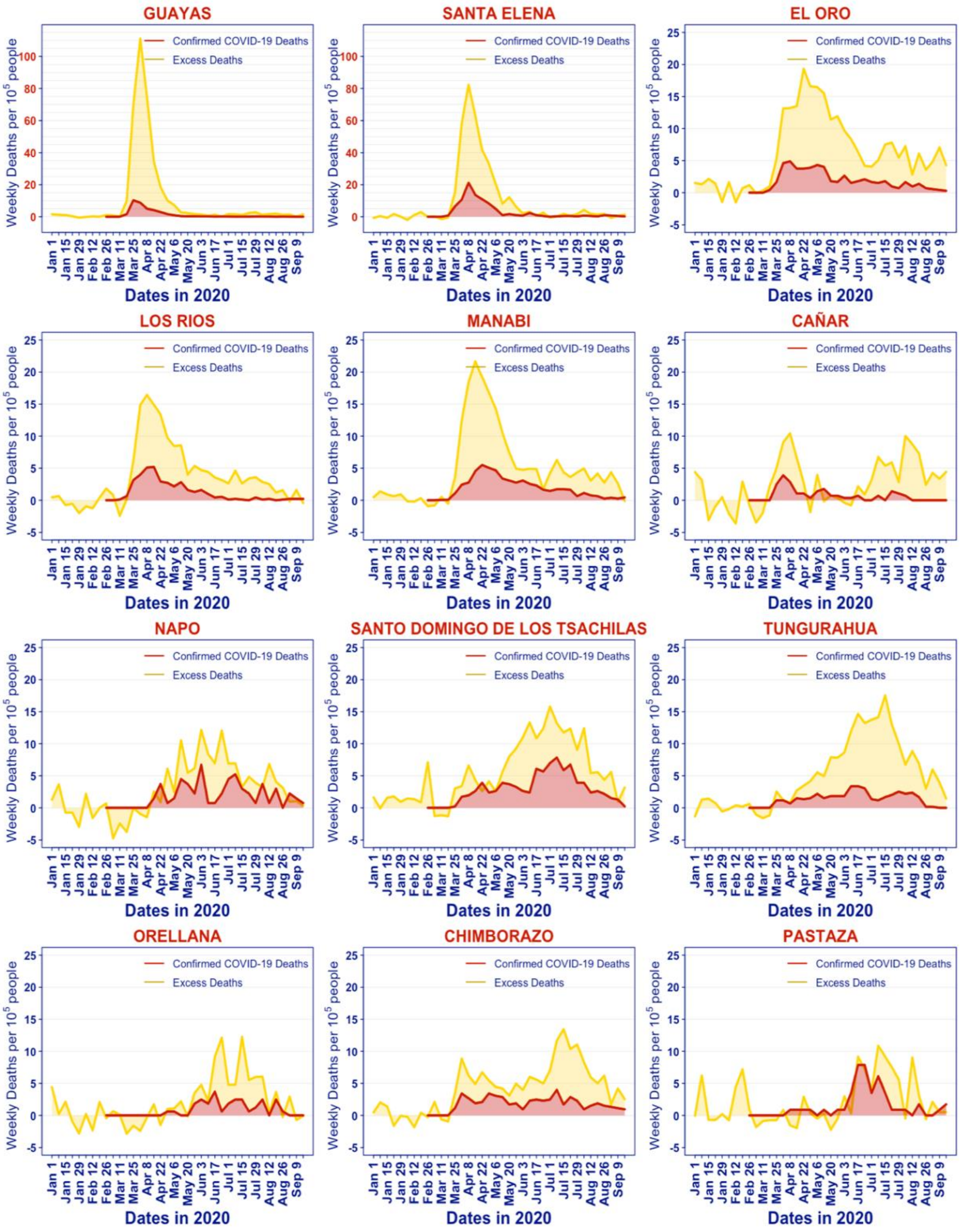


**Figure S5. Provincial time series of excess deaths per 100,000 people (yellow) and for documented COVID-19 deaths per 100,000 people (red).** Plots ordered according to the order in which provinces reached 10 excess deaths per 100,000 people. The first 12 provinces are shown.

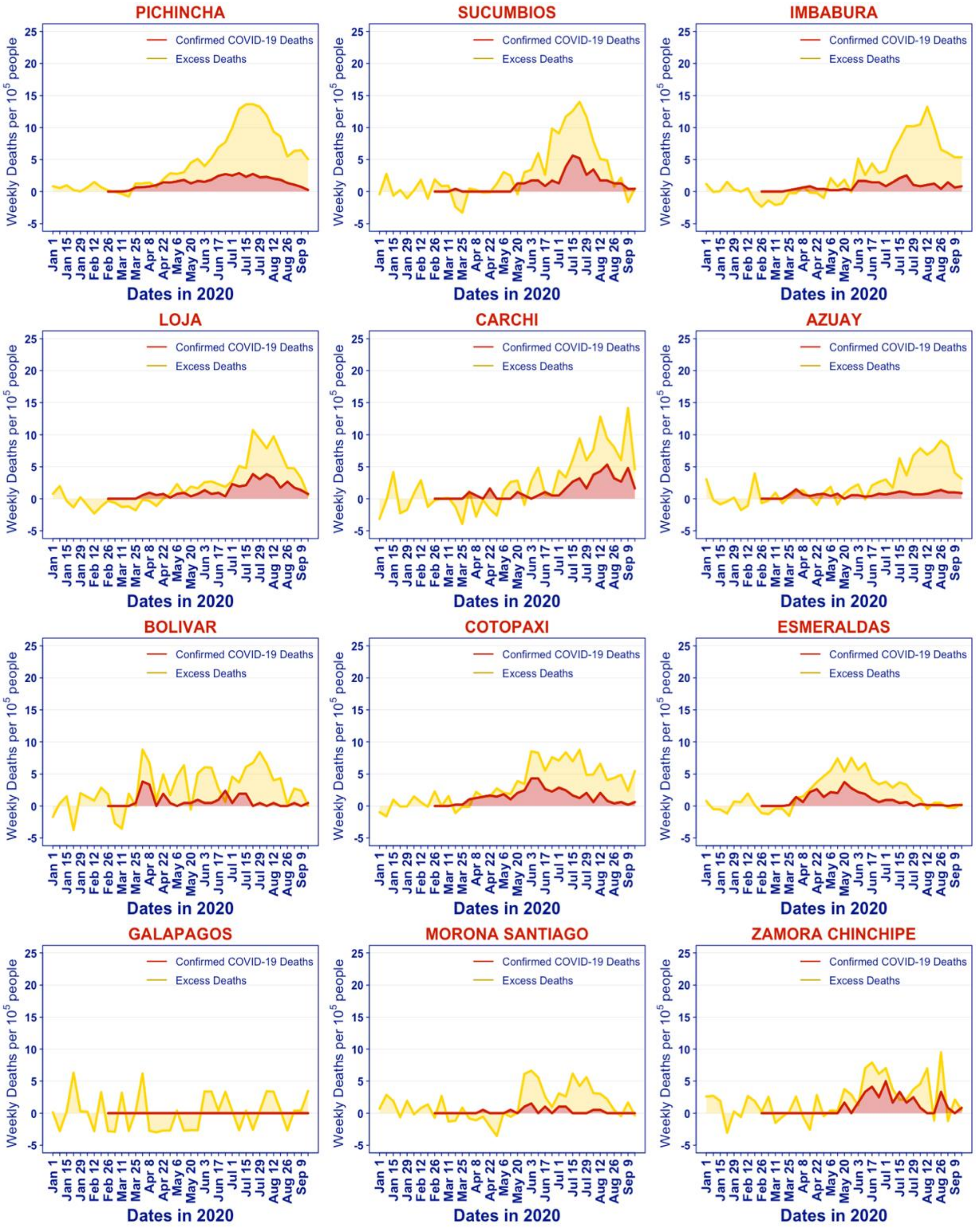


**Figure S6. Provincial time series of excess deaths per 100,000 people (yellow) and for documented COVID-19 deaths per 100,000 people (red).** Plots ordered according to the order in which provinces reached 10 excess deaths per 100,000 people. The last 12 provinces are shown
